# Comparison of the reactogenicity and immunogenicity between two-dose mRNA COVID-19 vaccine and inactivated followed by an mRNA vaccine in children aged 5 - 11 years

**DOI:** 10.1101/2022.11.07.22282028

**Authors:** Nasamon Wanlapakorn, Sitthichai Kanokudom, Harit Phowatthanasathian, Jira Chansaenroj, Nungruthai Suntronwong, Suvichada Assawakosri, Ritthideach Yorsaeng, Pornjarim Nilyanimit, Preeyaporn Vichaiwattana, Sirapa Klinfueng, Thanunrat Thongmee, Ratchadawan Aeemjinda, Nongkanok Khanarat, Donchida Srimuan, Thaksaporn Thatsanatorn, Natthinee Sudhinaraset, Yong Poovorawan

## Abstract

**Objective:** To compare the reactogenicity and immunogenicity between the two-dose mRNA COVID-19 vaccine regimen and one or two doses of inactivated vaccine followed by an mRNA vaccine regimen in healthy children between 5-11 years of age.

**Methods:** A prospective cohort study was performed at King Chulalongkorn Memorial Hospital in Thailand between March to June 2022. Healthy children between 5-11 years of age were enrolled and received the two-dose mRNA COVID-19 vaccine (BNT162b2) regimen or the inactivated (CoronaVac) vaccine followed by the BNT162b2 vaccine regimen. In addition, healthy children who received two doses of BBIBP-CorV between 1-3 months prior were enrolled to receive a heterologous BNT162b2 as a third dose (booster).

Reactogenicity was assessed by a self-reported online questionnaire. Immunogenicity analysis was performed to determine binding and surrogate neutralizing antibodies to SARS-CoV-2 wild-type and Omicron variants.

**Results:** Overall, 166 eligible children were enrolled. Local and systemic AE which occurred within 7 days after vaccination were mild to moderate and well-tolerated. At one-month, post-two or post-three doses, children vaccinated with two-dose BNT162b2, CoronaVac/BNT162b2, and two-dose BBIBP-CorV followed by BNT162b2 elicited similar levels of anti-receptor-binding domain (RBD) IgG. However, the two-dose BNT162b2 and two-dose BBIBP-CorV followed by BNT162b2 groups elicited higher neutralizing activities against Omicron BA.2 variant than the CoronaVac/BNT162b2 group.

**Conclusion:** The heterologous, CoronaVac vaccine followed by the BNT162b2 vaccine, regimen elicited lower neutralizing activities against the emerging Omicron BA.2 variant than the two-dose mRNA regimen. A third dose (booster) mRNA vaccine should be prioritized for this group.

## 1. Introduction

The COVID-19 pandemic, caused by SARS-CoV-2, has caused more than 600 million infections and more than 6.5 million deaths worldwide as of October 2022.^1^ The first COVID-19 vaccine that received emergency use authorization from the United States Food and Drug Administration (FDA) in December 2020 was the BNT162b2 vaccine for persons 16 years of age or older.^2, 3^ Later on, several effective COVID-19 vaccines were developed, tested, and initially distributed to the adult population, leaving children as a vulnerable population.

During the early COVID-19 pandemic, children infected with SARS-CoV-2 often developed mild symptoms.^4^ Nevertheless, the increase of symptomatic SARS-CoV-2 infection and hospitalization rate among children during the Delta (B.1.617.2) and Omicron (B.1.1.529) variants era indicated the need to extend vaccination to the pediatric population. Between October to December 2020, a phase 1/2 randomized controlled trial to assess the safety and immunogenicity of the inactivated CoronaVac vaccine in children between 3-17 years was conducted in China.^5^ The results showed that 3.0 µg CoronaVac was safe and able to elicit 100% seroconversion in children. In addition, another randomized controlled trial in 2020 demonstrated that the 4.0 µg inactivated BBIBP-CorV vaccine was safe for children 3-18 years and could elicit a robust humoral response after two doses.^6^

Apart from the inactivated vaccine, an mRNA vaccine has also been extended to the pediatric population. On October 29, 2021, the BNT162b2 vaccine was approved as an emergency use authorization for children 5-to-11-years old in the US based on the data in phase 2/3 trial which demonstrated that two doses of the 10 µg BNT162b2 vaccine administered 21 days apart showed an efficacy of 90.7% against the circulating SARS-CoV-2 variant.^2^ In December 2021, the Centers for Disease Control and Prevention (CDC) reviewed adverse events of the BNT162b2 vaccine from the Vaccine Adverse Event Reporting System (VAERS) and reported that myocarditis was associated with the mRNA-based COVID-19 vaccine.^7^ This rare but serious side effect raised concerns among the parents which could lead to vaccine hesitancy.

Thailand has secured the inactivated CoronaVac vaccine since February 2021, the inactivated BBIBP-CorV vaccine since June 2021, and the mRNA BNT162b2 vaccine since August 2021. In December 2021, the BNT162b2 vaccine was approved by the Thai FDA as a two-dose regimen for children aged between 5-11 years. In February 2022, the two-dose CoronaVac and two-dose BBIBP-CorV vaccines were approved by the Thai FDA for children aged between 6-11 years. The Ministry of Public Health and the Royal College of Thai Pediatricians recommended a dosing interval of 8 weeks for the two-dose BNT162b2 regimen based on an immunogenicity study reporting that a long-interval vaccination schedule induced higher antibody response compared to a short-interval vaccination schedule.^8^ Besides, the Ministry of Public Health and the Royal College of Thai Pediatricians also recommended that a heterologous inactivated (CoronaVac) vaccine followed by the BNT162b2 vaccine administered 4 weeks apart could be an alternative regimen for parents concerned about the adverse events following an mRNA vaccination.

Our previous immunogenicity study in Thai healthy adults vaccinated with a heterologous, CoronaVac vaccine followed by the BNT162b2 vaccine regimen showed that this regimen induced a higher antibody response compared to the homologous CoronaVac.^9^

Amidst the emergence of the Omicron variant, both two-dose mRNA or inactivated vaccination regimens cannot prevent breakthrough infections.^10, 11^ A third dose is recommended to obtain high immunity against the Omicron variant. Our previous studies in adults have shown that a booster mRNA vaccine after inactivated (CoronaVac or BBIBP-CorV)-primed individuals elicited a high level of antibody responses against the Omicron variant.^12, 13^ However, there was limited information about the safety and immunogenicity of a heterologous mRNA booster in inactivated vaccine-primed children.

This study aims to compare the reactogenicity and immunogenicity between the heterologous CoronaVac vaccine followed by the BNT162b2 vaccine regimen and the two-dose BNT162b2 vaccine regimen in children between 5-11 years of age. In addition, the reactogenicity and immunogenicity of the heterologous BNT162b2 booster in BBIBP-CorV-primed children were evaluated. The results of this study will help guide the physician’s decision on a mix-and-match vaccine strategy in certain circumstances and guide the booster strategy in the two-dose BBIBP-CorV-primed children.

## 2. Materials and methods

### 2.1. Study design and participants

This prospective cohort study was conducted between March to June 2022 at the clinical trial research unit at the Center of Excellence in Clinical Virology, Department of Pediatrics, Faculty of Medicine, Chulalongkorn University in Bangkok, Thailand. The study protocol was approved by the Institutional Review Board (IRB) of the Faculty of Medicine of Chulalongkorn University (IRB 059/65) and was performed under the principles of the Declaration of Helsinki. This trial was registered in the Thai Clinical Trials Registry (TCTR20220212001). Written informed consent was obtained from the parents or the legal guardians of participants prior to enrollment. Written assent was obtained from children aged 7 years and above. The inclusion criteria were immunocompetent children between 5-11 of age with no or well-controlled underlying diseases, no previous COVID-19 vaccination, and no previous SARS-CoV-2 infection from the medical history. The first group of participants was enrolled to receive the two-dose BNT162b2 regimen administered 8 weeks apart. The second group was enrolled to receive the CoronaVac followed by BNT162b2 vaccination administered 4 weeks apart. In addition, the third group of participants who had previously been immunized with two doses of the BBIBP-CorV vaccine between 1-3 months prior were enrolled. Participants received two doses of the BBIBP-CorV vaccine in other hospitals but need to provide immunization records in the electronic health record from the Ministry of Public health, Thailand prior to enrollment. This group is consented to receiving the third dose (booster) with BNT162b2. The inclusion criteria are the same as in the CoronaVac followed by BNT162b2 and homologous two-dose BNT162b2 groups, except the criteria of no previous COVID-19 vaccination.

### 2.2. Vaccine and blood collection

The CoronaVac vaccine from Sinovac Life Sciences, Beijing, China (hereafter referred to as SV) is an inactivated SARS-CoV-2 vaccine (CZ02 strain). The dosage for the pediatric population is 0.5 mL per dose containing 600 Spike Units (equal to 3 micrograms) of inactivated SARS-CoV-2 whole virus as antigen. The BBIBP-CorV vaccine from Sinopharm, Beijing, China (hereafter referred to as SP) is also an inactivated vaccine developed from the whole SARS-CoV-2 stain HB02. The interval recommended for the SP vaccine is two doses administered 4 weeks apart. The BNT162b2 from Pfizer-BioNTech, NY, USA (hereafter referred to as PF) is a lipid nanoparticle containing mRNA encoding the SARS-CoV-2 full-length spike of ancestral SARS-CoV-2 strain. The dosage for the pediatric population is 0.2 mL (10 micrograms) per dose. The recommended interval for SV/PF and PF/PF regimens are 4 and 8 weeks, respectively.

For participants in the SV/PF and PF/PF groups, blood samples were collected before the first dose vaccination (V1, baseline or pre-dose 1), before the second dose vaccination (V2, pre-dose 2), and one month after the second dose vaccination (V3, post-dose 2).

For participants in the SP/SP/PF group, blood samples were collected between 1-3 months after the second dose vaccination (V3, post-dose 2) and one month after the third dose vaccination (V4, post-dose 3).

### 2.3. Safety assessments

Parents or guardians of the participants recorded both local and systemic adverse events (AEs) after immunization within 7 days using self-administered online and paper questionnaires. An explanation of data collection was given to participants by trained investigators during the vaccination visit. Local and systemic AEs were classified as mild, moderate, and severe as previously described.^14^

### 2.4. Laboratory assessments

Serum samples were tested for binding antibody specific to the receptor-binding domain (RBD) of SARS-CoV-2, including total RBD immunoglobulin (Ig) and anti-RBD IgG, and anti-nucleocapsid (N) IgG as previously described.^15^ Neutralizing activities against wild-type (Euroimmun, Lubeck, Germany) and Omicron (BA.2) (GenScript Biotech, NJ, USA) were analyzed using a surrogate virus neutralization test (sVNT) as previously described.^16^ The seropositivity of sVNTs against wild-type and Omicron (BA.2) were determined as ≥35% and ≥30% inhibition, respectively. Samples were tested at the end of the study using the same batch of test kits.

### 2.5. Statistical analysis

The statistical differences in age between groups were performed using the Kruskal-Wallis test, followed by Dunn’s post hoc test with Bonferroni correction. Local and systemic adverse events between different regimens after the first and second doses were compared by risk differences with a 95% confidence interval (CI). Total RBD Ig and anti-RBD IgG were presented as geometric mean titers (GMT) with a 95% CI. Percent inhibitions by the sVNT assay were presented as a median with interquartile ranges (IQR). Differences in the geometric mean ratio (GMR) of total RBD Ig and anti-RBD IgG between groups were calculated by ANCOVA with Bonferroni’s adjustment. Differences in percentages of inhibition between groups were calculated by the Kruskal-Wallis test with Dunn’s multiple comparisons. A *p*-value of <0.05 was considered statistically significant.

## 3. Results

### 3.1 Demographic data and baseline characteristics

From March to June 2022, a total of 166 children were enrolled in the study. The consort flow diagram of study participants was shown in Figure 1. There were 43 eligible participants enrolled in SV/PF group. Among this group, there were 3 participants tested positive for COVID-19 after the first or second dose vaccination (breakthrough infection), 11 participants who had anti-N IgG, total RBD IgG, or anti-RBD IgG positive at baseline, presumably due to asymptomatic COVID-19 infection, and 2 participants who had evidence of SARS-CoV-2 infection as determined by anti-N IgG seroconversion. Therefore, a total of 16 participants in the SV/PF groups were excluded from the final immunogenicity analysis.

**Figure 1.**
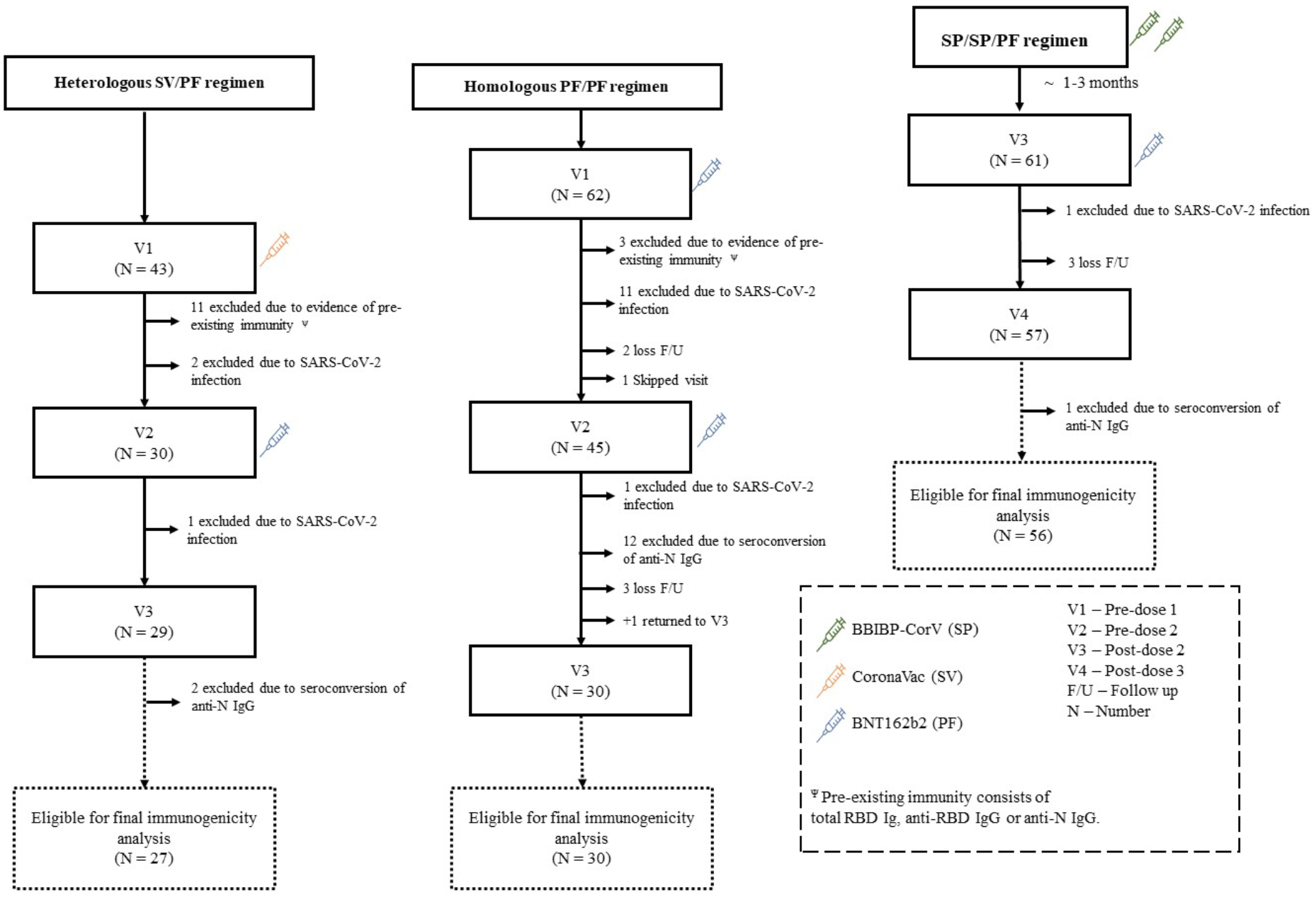
Study flow diagram of enrolled children (5-11 years) for final vaccine immunogenicity analysis. Unvaccinated children were enrolled to receive either heterologous CoronaVac/BNT162b2 (SV/PF) or homologous BNT162b2/BNT162b2 (PF/PF) regimen. In addition, the two-dose BBIBP-CorV-primed children were enrolled to receive the BNT162b2 as a third dose (SP/SP/PF). Sera were collected at pre-dose 1 (V1), pre-dose 2 (V2), post-dose 2 (V3), and post-dose 3 (V4), respectively. Children who contracted COVID-19 or had evidence of infection as determined by anti-N IgG seroconversion were excluded from the final vaccine immunogenicity analysis.

There were 62 eligible participants enrolled in PF/PF group. Among this group, there were 11 and 1 participant(s) tested positive for COVID-19 after the first and second dose vaccination, respectively (breakthrough infection), 3 participants who had anti-N IgG, total RBD IgG, or anti-RBD IgG positive at baseline, presumably due to asymptomatic COVID-19 infection, 12 participants who had evidence of SARS-CoV-2 infection as determined by anti-N IgG seroconversion, and 5 participants were lost to follow-up. Therefore, a total of 32 participants in the PF/PF groups were excluded from the final immunogenicity analysis.

There was 1 participant who skipped visit 2, received PF at a local hospital, and returned for blood testing on visit 3. There were 61 eligible participants enrolled in SP/SP/PF group. Among this group, there was 1 participant who tested positive for COVID-19 after the third dose of vaccination, 1 participant who had evidence of SARS-CoV-2 infection as determined by anti-N IgG seroconversion, and 3 participants who were lost to follow-up. Therefore, a total of 5 participants in the SP/SP/PF group were excluded for the final immunogenicity analysis.

The demographics and characteristics of the participants are shown in Table 1. There were 27, 30, and 56 participants in the SV/PF, PF/PF, and SP/SP/PF groups who were included in the final immunogenicity analysis, respectively. The number of female participants per total (%) participants were similar among groups. Nevertheless, there was a statistically significant difference in age between the PF/PF group (mean 6.2 years) and SP/SP/PF group (mean 7.8 years) (P-value <0.001). The mean interval between doses 1 and 2 were 30.2, 57.4, and 22.9 days in the SV/PF, PF/PF, and SP/SP/PF groups, respectively. The mean interval between doses 2 and 3 was 61.0 days in the SP/SP/PF group.

**Table 1.** Demographics and characteristics of the enrolled children (5-11 years).

| Group | Participants eligible for final immunogenicity analysis |  |  | Participants who had pre-existing antibody | Participants who had seroconversion of anti-N IgG |
| --- | --- | --- | --- | --- | --- |
|  | SV/PF | PF/PF | SP/SP/PF | SV/PF ¥ | PF/PF § |
| **Total** | 27 | 30 | 56 | 11 | 12 |
| **Sex, Female/total (%)** | 12/27 (44.4) | 16/30 (53.3) | 27/56 (48.2) | 6/11 (54.5) | 10/12 (83.3) |
| **Mean age in year (SD)** | 7.4 (2.2) | 6.2 (1.1) | 7.8 (1.6) | 6.3 (1.6) | 6.8 (1.8) |
| **No comorbidity (%)** | 24/27 (88.9) | 27/30 (90.0) | 48/56 (85.7) | 11/11 (100) | 12/12 (100) |
| **Underlying diseases (%)** |  |  |  |  |  |
| **Allergy** | 2/27 (7.4) | 2/30 (6.7) | 6/56 (10.7) | - | - |
| **Asthma** | - | 1/30 (3.3) | - | - | - |
| **Autism spectrum disorder** | - | - | 1/56 (1.8) | - | - |
| **Obstructive sleep apnea** | 1/27 (3.7) | - | - | - | - |
| **Thalassemia trait** | - | - | 1/56 (1.8) | - | - |
| **Time interval between first and second dose** | 30.2 (28-41) | 57.4 (57-67) | 22.9 (21-82) | 58.0 (57-67) | 58.2 (57-67) |
| **Mean (range), days** |  |  |  |  |  |
| **Time interval between second dose and blood sampling** | 29.3 (27-37) | 31.0 (24-46) | - | 32.0 (27-36) | 32.0 (27-36) |
| **Mean (range), days** |  |  |  |  |  |
| **Time interval between second dose and third dose** | - | - | 61.0 (27-167) | - | - |
| **Mean (range), days** |  |  |  |  |  |
| **Time interval between third dose and blood sampling** | - | - | 31.0 (29-36) | - | - |
| **Median (range), days** |  |  |  |  |  |

¥ refers to the children who had pre-existing antibody before receipt of 1<sup>st</sup> dose of CoronaVac.
§ refers to the children who had seroconversion of anti-N IgG before receipt of 2<sup>nd</sup> dose of BNT162b

We also compared the antibody responses elicited by the different vaccine regimens with those elicited by vaccine regimens plus natural infection (hybrid immunity). Hybrid immunity groups refer to participants who had anti-N IgG seroconversion after receiving the vaccine or participants who had pre-existing antibodies (anti-N IgG or total RBD IgG) at baseline before the first vaccine as described in Table 1.

### 3.2 Safety and reactogenicity profile

The most common solicited local adverse reaction (AE) after vaccination was pain at the injection site: SV/PF group (first dose 62.5%; second dose 68.6%), PF/PF group (first dose 59.3%; second dose 57.8%), SP/SP/PF group (third dose; 85.2%). The most common systemic AE was myalgia (20.0-28.6%) (Figure S1). Comparisons of AEs between SV/PF and PF/PF regimens after the first or second dose vaccination showed no significant differences in any of the local or systemic AEs (Figure 2). Most of the solicited local and systemic AEs were mild (grade 1) or moderate (grade 2) and resolved within a few days post-vaccination (Figure S1). Frequencies of grade 3 local or systemic AEs after ranged between 1% to 3%. No serious AEs were reported.

**Figure 2.**
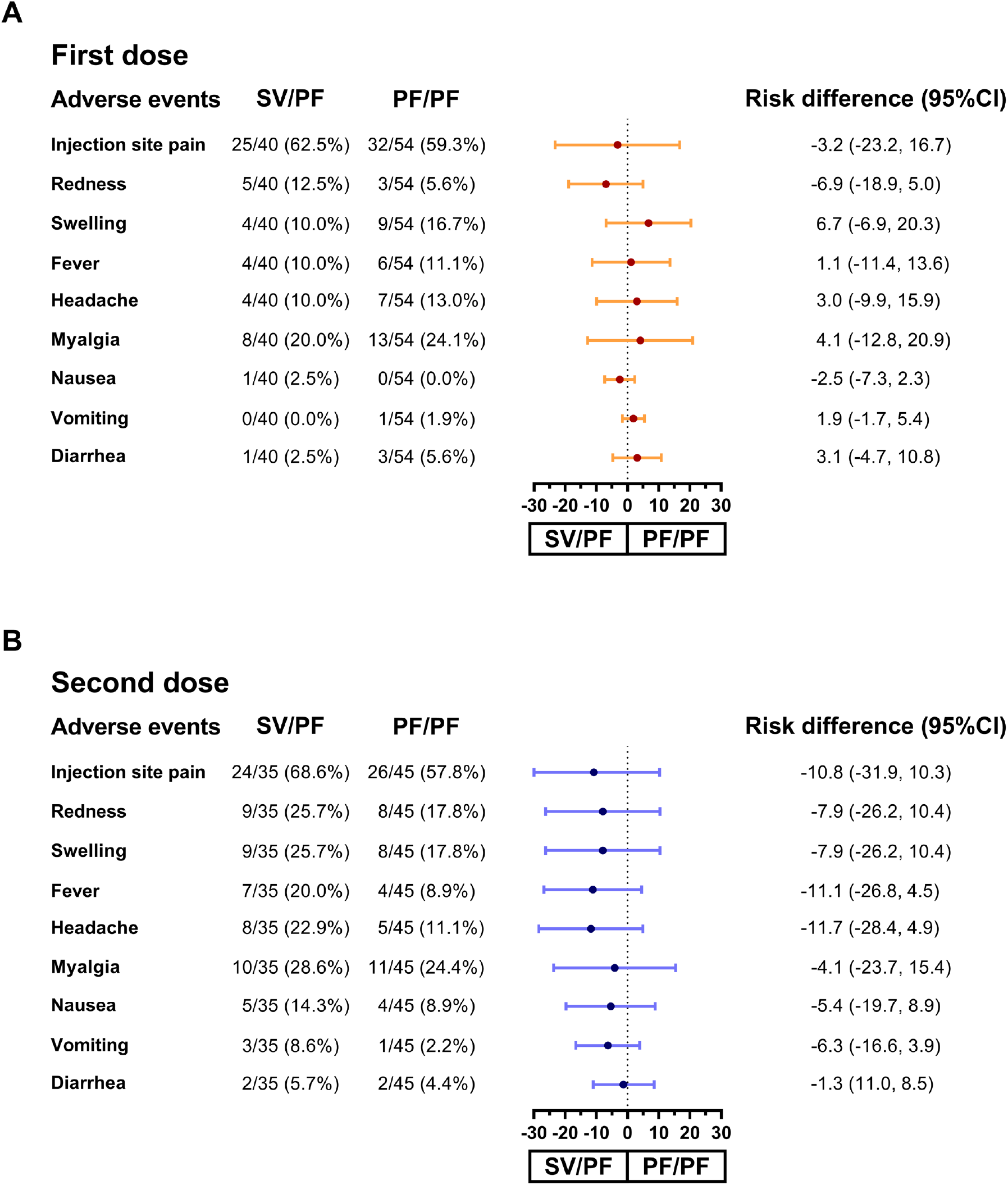
Forest plot showing the percentages of solicited local and systemic adverse events (AEs) and the risk differences with 95% confidence intervals (95% CI) in pediatric participants with any grade AEs across 7 days after vaccination. The AEs of first dose vaccination (A) and second dose vaccination (B) between heterologous SV/PF regimen and homologous PF/PF regimen were compared. SV and PF refer to CoronaVac and BNT162b2, respectively.

### 3.3 Total RBD Ig and anti-RBD IgG responses in pediatric participants after different regimens of COVID-19 vaccines

The geometric mean titer (GMT) of total RBD immunoglobulin (Ig) which predominantly included IgG, but also some amount of IgM and IgA, and anti-RBD IgG were compared among groups vaccinated with different regimens using ANCOVA with Bonferroni’s adjustment as shown in Figure 3. There were no differences in total RBD Ig and anti-RBD IgG at pre-vaccination among the SV/PF and PF/PF groups at pre-dose 1 (V1). At one-month post-dose 2 (V3), the PF/PF group had significantly higher total RBD Ig than the SV/PF group (Fig 3A) (geometric mean ratio (GMR) 2.04). However, there were no differences in anti-RBD IgG levels among the SV/PF and PF/PF groups (Fig 3B). In addition, the three-dose SP/SP/PF group also possessed higher total RBD Ig than the two-dose SV/PF and PF/PF groups (GMR 3.09 and 1.52, respectively), but there was no difference in the anti-RBD IgG levels among all groups.

**Figure 3.**
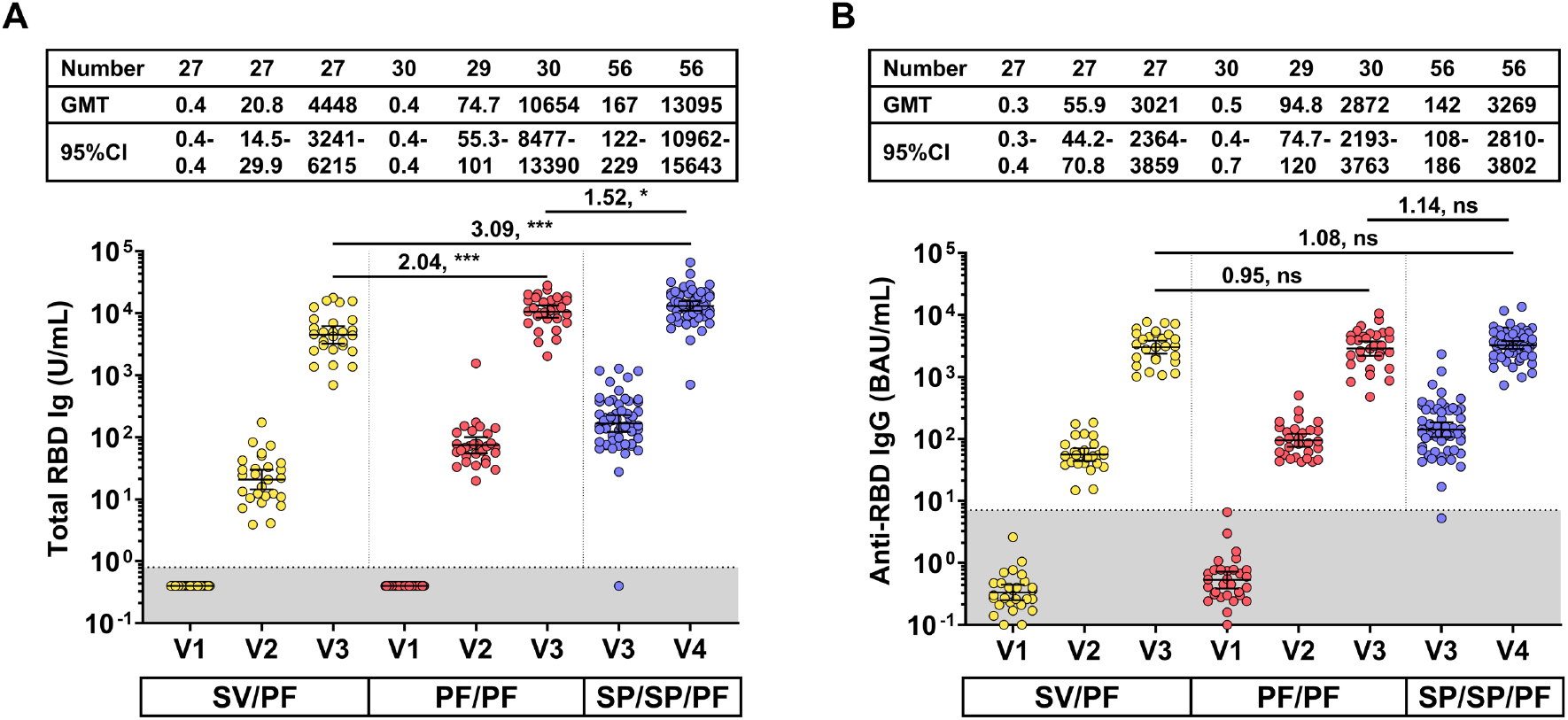
Binding antibody specific for SARS-CoV-2 in vaccinated children (5-11 years). A) Total immunoglobulin specific to the receptor-binding domain (RBD) (Total RBD Ig) (U/mL) and B) anti-RBD IgG (BAU/mL) of the heterologous CoronaVac/BNT1612b2 (referred to as SV/PF), homologous BNT1612b2 (referred to as PF/PF), and third dose of BNT162b2 in two-dose BBIBP-CorV-primed (refers to SP/SP/PF) groups. For both graphs the intervals of reported immunological values are as follows: on the day of the first dose (V1; pre-dose 1), 4 and 8 weeks later for SV/PF and PF/PF groups, respectively (V2; pre-dose 2), 4 weeks after two-dose completion in the SV/PF and PF/PF groups and 1-3 months after two-dose completion in the SP/SP/PF group (V3; post-dose 2), and 4 weeks after three-dose completion in the SP/SP/PF group (V4; Post-dose 3). Data points are the reciprocals of the individual. The gray area indicates the seronegativity of total RBD Ig (<0.8 U/mL) or anti-RBG IgG (<7.1 BAU/mL). Lines indicate geometric means and bars indicate 95% confidence intervals (95%CI). A pairwise comparisons display geometric mean ratio (GMR) and significant values including p <0.05 (*), p <0.001 (***) and no statistical significance (ns).

### 3.4 Total RBD Ig and anti-RBD IgG responses between two-dose vaccination and hybrid immunity

There was a subgroup of participants (n=11) in the SV/PF group found to have a pre-existing anti-N IgG, total RBD IgG, or anti-RBD IgG at baseline (before receiving the first dose SV). The baseline characteristics of this subgroup was shown in Table 1. This group was excluded from the immunogenicity analysis as they presumably had asymptomatic or unrecognized COVID-19 infection prior to enrollment. However, all the immunogenicity results were available at the end of the study and children in this group had received the two-dose SV/PF vaccine. Therefore, antibody levels in this SV/PF-vaccinated group who possessed pre-existing immunity (refers to as pre-existing immunity group) were compared to the SV/PF-vaccinated group without pre-existing immunity as shown in Figures 4A and B. The results showed that the pre-existing immunity group elicited higher total RBD Ig at pre-dose 2 (V2) and post-dose 2 (V3) than those without pre-existing immunity. However, the anti-RBD IgG at post-dose 2 (V3) was comparable between both groups.

**Figure 4.**
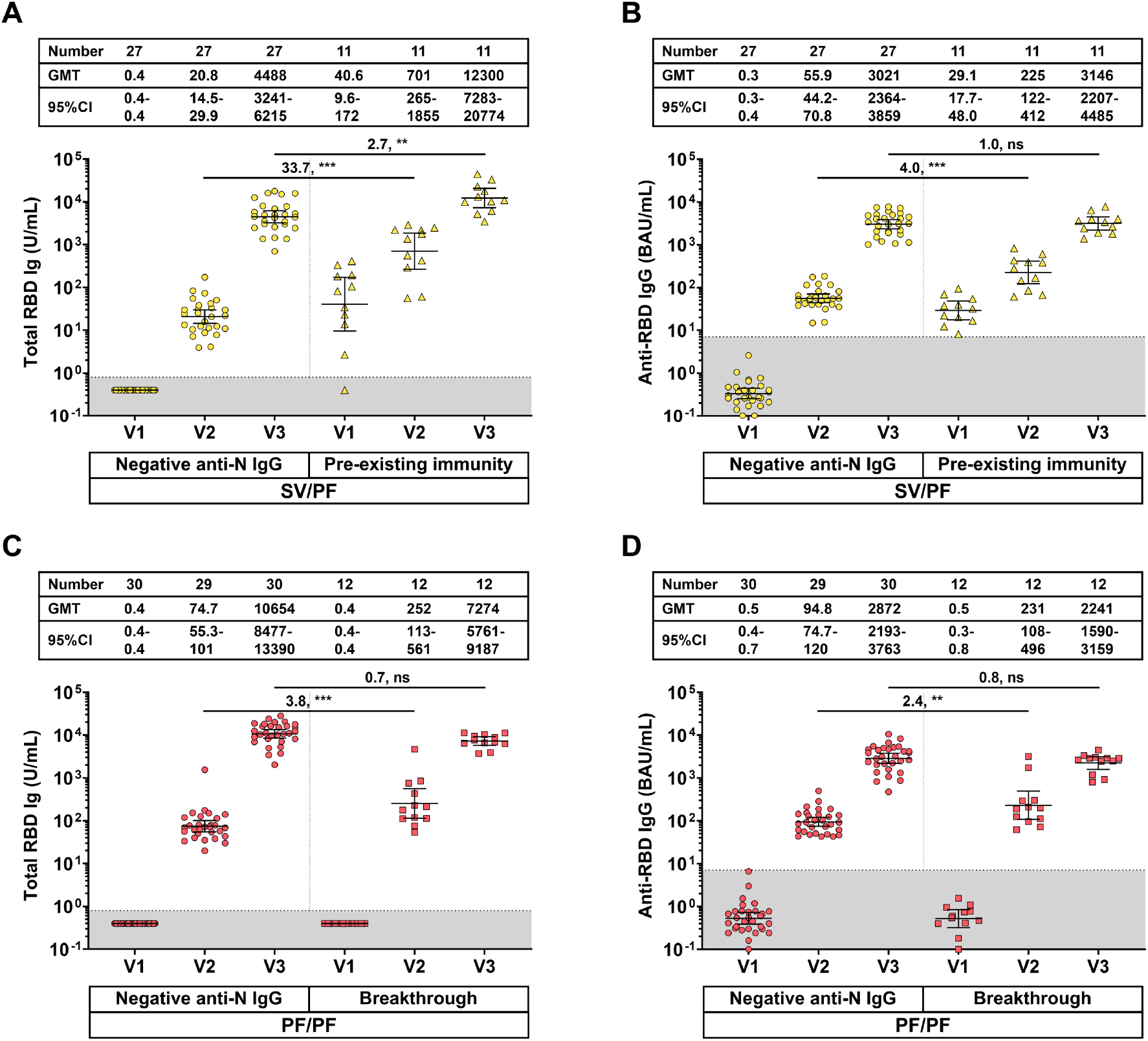
Binding antibody specific for SARS-CoV-2 classified by immunity status. A) Total immunoglobulin specific to the receptor-binding domain (RBD) (Total RBD Ig) (U/mL) and B) anti-RBD IgG (BAU/mL) of the heterologous CoronaVac/BNT1612b2 (referred to as SV/PF) group classified by presence or absence of pre-existing immunity at baseline. C) Total RBD Ig (U/mL) and D) Anti-RBD IgG (BAU/mL) of the homologous BNT1612b2 (referred to as PF/PF) group classified by breakthrough or non-breakthrough infection. For all graphs the intervals of reported immunological values are as follows: on the day of the first dose (V1; pre-dose 1), 4 and 8 weeks later for SV/PF and PF/PF groups, respectively, (V2; pre-dose 2), and 4 weeks after two-dose completion (V3; post-dose 2). Data points are the reciprocals of the individual. The gray area indicates the seronegativity of total RBD Ig (<0.8 U/mL) or anti-RBG IgG (<7.1 BAU/mL). Lines indicate geometric means and bars indicate 95% confidence intervals (95%CI). A pairwise comparisons display geometric mean ratio (GMR) and significant values including p <0.01 (**), p <0.001 (***) and no statistical significance (ns).

Similarly, there was a subgroup of participants (n=12) in the PF/PF group found to have been infected with SARS-CoV-2 between vaccination visits 1 and 2 as evidenced by the seroconversion of anti-N IgG. The baseline characteristics of this subgroup was shown in Table 1. This group was excluded from the immunogenicity analysis as they presumably had asymptomatic or unrecognized COVID-19 infection after receiving the first dose of the BNT162b2 vaccine. Similarly, all the immunogenicity results were available at the end of the study and children in this group had received the second dose BNT162b2 vaccine. Antibody levels in this PF/PF-vaccinated group who had a breakthrough infection (refers to as the breakthrough group) were compared to the PF/PF-vaccinated group without breakthrough infection as shown in Figure 4C and D. The results showed that the breakthrough group possessed higher levels of total RBD Ig and anti-RBD IgG than the non-breakthrough group at pre-dose 2 but not at post-dose 2.

### 3.5 Neutralizing activity against wild-type and Omicron BA.2 using sVNT

Neutralizing activities against wild-type SARS-CoV-2 after two-dose or three-dose vaccination was above 99% and similar among all groups (Figure 5A). However, following the second dose vaccination, the homologous PF/PF group possessed higher neutralizing activities against the Omicron BA.2 variant compared to the heterologous SV/PF group (p < 0.001) (Figure 5B). In addition, the three-dose vaccinated group (SP/SP/PF) also elicited higher neutralizing activities against the Omicron BA.2 variant compared to the heterologous SV/PF group (p < 0.001).

**Figure 5.**
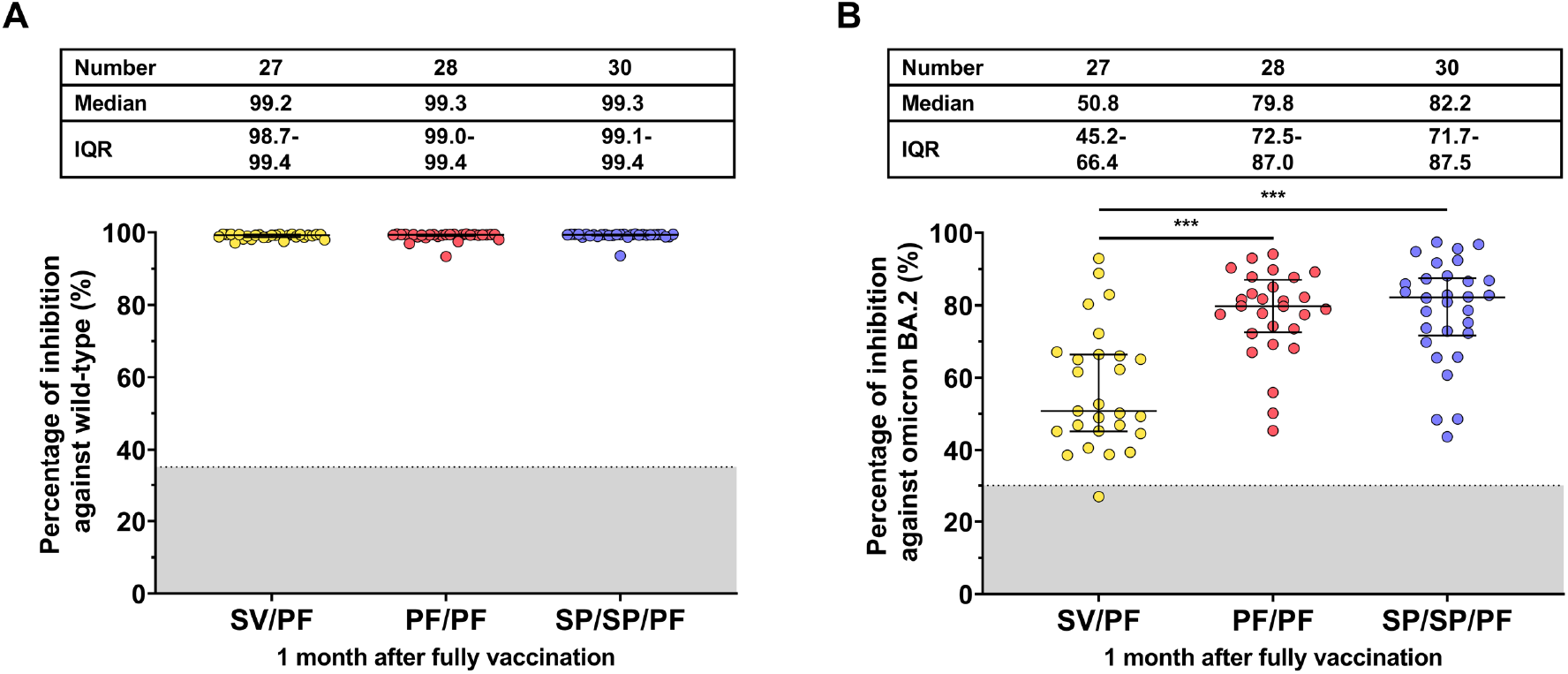
Neutralizing activity against the A) wild-type and B) BA.2 Omicron variant at one month after a two-dose or three-dose vaccination. Lines indicate median percent inhibition and the error bar indicates the interquartile range (IQR). The gray area indicates the seronegativity of neutralizing activity of the wild-type (<35%) and BA.2 Omicron variant (<30%). A pairwise comparisons display statistical significance, p <0.001 (***).

## 4. Discussion

In this study, we evaluated the extent of binding and neutralizing antibody responses to the two-dose BNT162b2 regimen and the heterologous CoronaVac followed by the BNT162b2 vaccine regimen in children between 5-11 years. In addition, we also evaluated the antibody response after a BNT162b2 vaccine as a booster dose in pediatric individuals who had received two-dose vaccination (priming) with the inactivated BBIBP-CorV vaccine regimen. Our study found that at one month post two-dose or three-dose vaccination, children vaccinated with the two-dose BNT162b2 regimen, the heterologous regimen, and the two-dose BBIBP-CorV followed by BNT162b2 regimen elicited similar levels of anti-RBD IgG. All vaccinated groups elicited neutralizing activities against wild-type and Omicron BA.2 variants of SARS-CoV-2 after completion of two-dose or three-dose vaccination. However, neutralizing activities as determined by percent inhibitions against the Omicron BA.2 variant were lower than that toward the ancestral strain. Furthermore, the two-dose BNT162b2 and two-dose BBIBP-CorV followed by BNT162b2 groups elicited higher neutralizing activities against Omicron BA.2 variant than the CoronaVac/BNT162b2 group.

Regarding the two-dose vaccine regimen in children, the real-world effectiveness of the two-dose CoronaVac or BNT162b2 vaccine regimen amidst the Omicron variant outbreak showed comparable results. A study in Chile demonstrated that the effectiveness of the two-dose CoronaVac regimen in children 3-5 years of age was 38.2% against symptomatic COVID-19, 64.6% against hospitalization, and 69.0% against ICU admission.^17^ Similarly, vaccine effectiveness against hospitalization during the Omicron predominance in children 5 to 11 years old elicited by two doses of BNT162b2 was 68%.^18^ Although the present study did not evaluate the effectiveness, the immunogenicity results showed that neutralizing activities against Omicron BA.2 variant and total RBD Ig in the two-doses BNT162b2 group administered 8 weeks apart were higher than the heterologous CoronaVac/BNT162b2 administered at 4 weeks apart. Previous studies on adenoviral-vectored and mRNA COVID-19 vaccine showed that increased dosing intervals improved the vaccine immunogenicity and effectiveness. ^8, 19^ Similarly, the extended dosing interval of the CoronaVac/BNT162b2 mix-and-match strategy also showed improved immunogenicity in adults.^9^ In Thailand, two doses of the BNT162b2 vaccine were administered 8 weeks apart, instead of 3 weeks apart as recommended by the Advisory Committee on Immunization Practices (ACIP),^20^ due to the limited vaccine supply and improved immunogenicity. Nevertheless, the CoronaVac/BNT162b2 regimen administered 4 weeks apart was recommended. Due to the lower immunogenicity profile of CoronaVac/BNT162b2, it is worth noting that the CoronaVac/BNT162b2-vaccinated group should be prioritized for the third dose booster.

Several studies in adults showed that heterologous boosters (mRNA vaccine as the third dose in inactivated COVID-19 vaccine-primed individuals) could enhance antibody response against the emerging Omicron variant.^12, 21^ In this study, we evaluated the immunogenicity after the BNT162b2 booster in BBIBP-CorV-primed individuals 1-3 months prior and found that the boosted children can elicit neutralizing antibody response against Omicron BA.2 variant similar to those elicited by two-dose BNT162b2. Our previous study found that longer interval between primary doses of CoronaVac and booster (i.e., 6 months) could enhance the total Ig and anti-RBD IgG responses compared to the short interval booster (i.e., 3 months).^12^ Nevertheless, the present study chose the short interval for BBIBP-CorV-primed individuals to receive a booster because Omicron-specific anti-RBD IgG elicited by the two-dose inactivated vaccine were low at 3 months after second dose, and get boosted significantly after an mRNA vaccine as the third (booster) dose.^21^ Thus, the heterologous mRNA booster in children primed with inactivated vaccine regimen should be recommended to increase protection against the emerging Omicron variant. although a longer interval between the second and the third dose could stimulate higher antibody responses.

All COVID-19 vaccines administered as primary or booster doses in this study had acceptable reactogenicity with mild to moderate AEs that generally resolved within a few days after vaccination. The adverse events reported herein were similar to those of pain, myalgia, and fever described in the previous studies.^2, 22^

Nearly one-third of the participants in the CoronaVac/BNT162b2 group had been exposed to the SARS-CoV-2 virus prior to enrollment. This was detected by seropositivity of anti-nucleocapsid (N) IgG or total RBD Ig at baseline. This study showed that total RBD Ig are higher in previously infected vaccinees than in SARS-CoV-2 naïve subjects following the first dose of the CoronaVac vaccination and at one month post-second dose (BNT162b2) vaccination. Our findings are in line with a study in adults showing that previously infected individuals who received a booster COVID-19 vaccine elicited higher neutralizing antibodies compared to the infection alone or vaccine alone.^23^ Regarding the two-dose BNT162b2 groups, there was a long waiting time (8 weeks) between the first and second doses of the BNT162b2 vaccine. Thus, nearly one-third of the participants in the two-dose BNT162b2 group had breakthrough infection after receipt of the first dose of the BNT162b2 vaccine as determined by seroconversion of anti-N IgG. Similarly, at visit 2 (post one dose + natural infection), breakthrough infection vaccinees had anti-RBD IgG and total RBD Ig higher than in SARS-CoV-2 naïve subjects immunized with one-dose BNT162b2 vaccine, but this difference disappeared after the second dose vaccine. This is also in agreement with a previous study in adults which showed that previously infected vaccinees who received 2 doses mRNA vaccine elicited higher Omicron BA.1-specific neutralizing antibodies similar to the triple-vaccinated subjects, but higher than SARS-CoV-2 naïve subjects who received 2 doses mRNA vaccine.^24^

Potential limitations of our study may be attributed to the loss of some participants from the final immunogenicity analysis due to the peak of the SRAS-CoV-2 outbreak in Bangkok during March to April 2022. The BNT162b2 vaccine was in multiple-dose vials which needed to be administered in a short period. The time limit of keeping opened multi-dose vials made the randomization not feasible in our study. As of May 2022, there was a recommendation from the Centers for Disease Control and Prevention that children who receive two-dose BNT162b2 should get a BNT162b2 booster. Thus, additional studies on the immunogenicity and efficacy of the third dose (booster) in two-dose BNT162b2 or heterologous CoronaVac/BNT162b2-primed children should be further investigated.

## 5. Conclusions

Local and systemic AE which occurred within 7 days after vaccination were mild to moderate and well-tolerated. The heterologous, CoronaVac vaccine followed by the BNT162b2 vaccine, regimen elicited lower neutralizing activities against the emerging Omicron BA.2 variant than the two-dose mRNA regimen. A third dose (booster) mRNA vaccine should be prioritized for this group.

## Data Availability

All data produced in the present work are contained in the manuscript

## Acknowledgments

We would like to thank all Center of Excellence in Clinical Virology personnel and all participants for contributing to and supporting this project. This research was financially supported by the Health Systems Research Institute (HSRI), National Research Council of Thailand (NRCT), the Center of Excellence in Clinical Virology, Chulalongkorn University, and King Chulalongkorn Memorial Hospital, and partially supported by the Second Century Fund (C2F) of Sitthichai Kanokudom, Chulalongkorn University.

## Author Contributions

Conceptualization, N.W., S.K. (Sitthichai Kanokudom), N.S.(Nungruthai Suntronwong), P.N. and Y.P.; data curation, N.W., S.K. (Sitthichai Kanokudom), N.S. (Nungruthai Suntronwong), S.A., R.Y., D.S., T.T. (Thaksaporn Thatsanatorn) and N.S. (Natthinee Sudhinaraset); formal analysis, N.W., S.K. (Sitthichai Kanokudom) and H.P.; methodology, S.K. (Sitthichai Kanokudom), J.C., P.V., S.K. (Sirapa Klinfueng), T.T (Thanunrat Thongmee), R.A. and N.K.; project administration, Y.P.; writing—original draft, N.W., S.K. (Sitthichai Kanokudom); writing—review and editing, N.W., S.K. (Sitthichai Kanokudom) and Y.P. All authors have read and agreed to the published version of the manuscript.

## Institutional Review Board Statement

The study protocol was approved by the Institutional Review Board (IRB), Faculty of Medicine, Chulalongkorn University (IRB number 059/65). This trial was registered in the Thai Clinical Trials Registry (TCTR20220212001).

## Informed Consent Statement

Informed consent was obtained from parents or legal guardians before participant enrollment. The study was conducted according to the Declaration of Helsinki and the Good Clinical Practice Guidelines (ICH-GCP) principles.

## Data Availability Statement

The datasets generated and analyzed during the current study are available from the corresponding author upon reasonable request.

