## Supplementary material for "Comparison of the reactogenicity and immunogenicity between two-dose mRNA COVID-19 vaccine and inactivated followed by an mRNA vaccine in children aged 5 - 11 years": Supplementary Figure_Homologous and heterologous COVID-19 vaccine in children.docx

**Supplementary Figure**
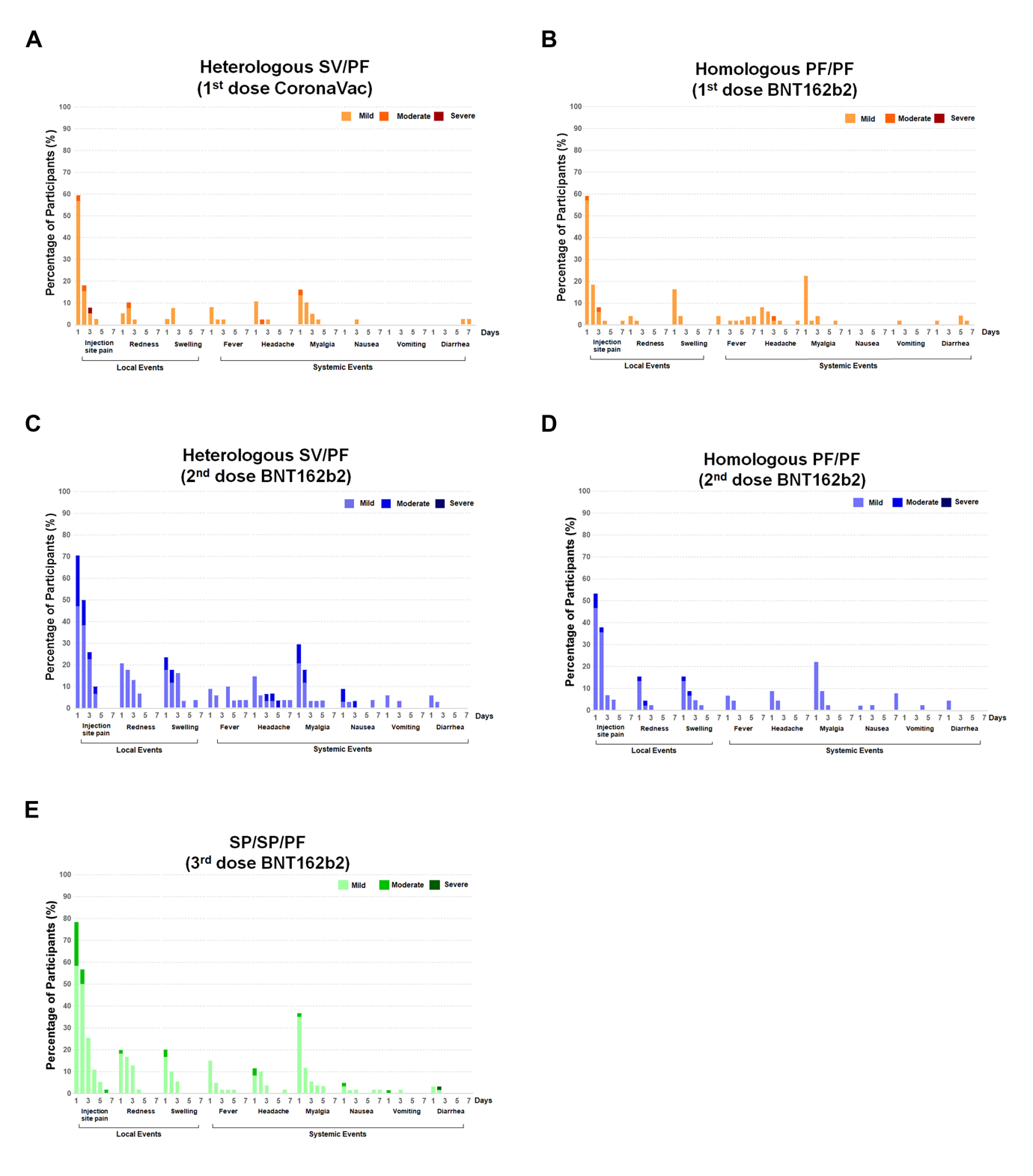
**Supplementary Figure 1.** Reactogenicity of pediatric participants after receiving one dose, two doses, or three doses of theCOVID-19 vaccine. Local and systemic adverse events were recorded 7 days post-vaccination. Reactogenicity of the (A) first dose of CoronaVac in the SV/PF group, (B) first dose of BNT162b2 in the PF/PF group, (C) second dose of BNT162b2 in the SV/PF group, (D) second dose of BNT162b2 in the PF/PF group, and (E) third dose of BNT162b2 in the SP/SP/PF groups. Local and systemic symptoms were graded as mild (easily tolerated with no limitation on regular activity), moderate (some limitation of daily activity), and severe (unable to perform regular daily activity).
